# The Representativeness of Regional Influenza Virus Genomic Surveillance for National Trends in the United States

**DOI:** 10.64898/2026.02.23.26346422

**Authors:** Manon Ragonnet-Cronin, Leigh Papalambros, Emily E. Bendall, William J. Fitzsimmons, Christopher N. Blair, Robert Tibbetts, Ashish Bhargava, Adam Lauring

## Abstract

Genomic surveillance of influenza viruses informs vaccine strain selection and evolutionary forecasting. Sequencing efforts vary widely across U.S. states, which raises concerns about spatial sampling bias. We evaluated how well 10,958 influenza virus genomes sampled by our group in Michigan captured the genetic diversity in 34,743 genomes circulating nationally from the 2021/22 through 2024/25 seasons. We defined seasonal hemagglutinin haplotypes and tracked their detection across states. A small number of haplotypes dominated each season, and Michigan detected all major haplotypes, even under substantial downsampling. Detection delays were primarily driven by haplotype frequency rather than geographic factors. Comparisons across states showed that higher sequencing effort improved coverage and detection timeliness, with diminishing returns at higher volumes. Rarefaction analysis confirmed that relatively few sequences were needed to capture 95% of national haplotype diversity. These findings suggest that intensive sequencing in a single well-sampled location can be broadly representative of national influenza diversity.

**One sentence summary:** Dense influenza genomic sequencing from a single U.S. state captured nearly all nationally circulating haplotype diversity, with detection timeliness primarily driven by sequencing effort and haplotype frequency.

## Introduction

Genomic surveillance has become increasingly central to infectious disease monitoring. Influenza remains a serious threat to public health, and sequence data have long been critical for vaccine strain selection, allowing for early identification of antigenically drifted lineages [1].

The COVID-19 pandemic further demonstrated the value of real-time genetic surveillance for characterizing emerging viral mutations, tracking variant or clade growth, and informing short-term forecasts of viral evolution and epidemic dynamics [2]. Rapid identification of novel and fast-growing strains of SARS-CoV-2 or influenza can signal low population immunity and forecast potential increases in infection burden. Models developed to make regular SARS-CoV-2 variant frequency forecasts based on sequence data [3] have now been extended to produce influenza clade frequency forecasts [4,5]. In addition, phylodynamic analyses of influenza viruses can be used to estimate epidemiological parameters and further uncover detailed transmission dynamics [6,7].

In the United States (U.S.), influenza surveillance at the national level is coordinated by the Centers for Disease Control (CDC), but viruses are collected, and often sequenced, by the states themselves [8]. Since the COVID-19 pandemic, many academic and commercial laboratories sample viruses and contribute sequences to public databases. However, sequencing is expensive and requires technical expertise to carry out strategically. Representative, random samples are ideal for making unbiased downstream inferences about viral evolution, fitness, and antigenic properties; but in many states, convenience samples are collected, or sampling strategies are undocumented. The range in sequencing efforts, sampling strategies and timeliness creates the possibility of spatial sampling bias.

Since 2021, the University of Michigan (UOM) has conducted deep, random sampling at locations across the state of Michigan. Here, we evaluate how well influenza virus genomes sampled in Michigan capture the genetic diversity of viruses circulating across the broader U.S. during the influenza seasons 2021/22 to 2024/25. By quantifying representativeness at the state level, this analysis provides an empirical check on a common practical assumption in downstream modeling and evolutionary analysis: that a dense sequence set from one well-sampled area can stand in for the entire country.

## Methods

### Specimen Sources

The UOM surveillance system captured nearly all test-positive respiratory specimens from testing laboratories affiliated with three large health systems - Michigan Medicine, Henry Ford Health System, and Ascension St. John (subsequently renamed Henry Ford Ascension) from October 2021 to September 2025. Together, these health systems provide the majority of care to residents of Michigan Response Region 2, which includes Wayne, Oakland, Macomb, and Washtenaw Counties (>4.5 million residents). These centralized laboratories performed testing for inpatients and outpatients who sought medical attention for a presumed respiratory illness.

### Sequencing

For isolates sequenced at UOM, nucleic acid was extracted using the MagMAX Viral/Pathogen II Nucleic Acid Isolation Kit (ThermoFisher) on a KingFisher instrument. The 8 genomic segments of influenza A virus were amplified in a one-step multiplex reverse transcription polymerase chain reaction (RT-PCR) [9]. Amplified cDNA were prepared for sequencing using either the Oxford Nanopore Technologies (ONT) or Illumina platforms. For ONT, unique barcodes were ligated to RT-PCR products from each sample using end repair and ligation kits (New England Biolabs) and then pooled in batches of 96. These pools were ligated to pore adaptors and sequenced with a GridION instrument. For Illumina, libraries were prepared using the Illumina DNA Prep Tagmentation Kit and sequenced in batches of 192 on a MiSeq instrument with 2×250 PE reads, v2 chemistry or 384 on a NextSeq 1000 instrument with 2×300 PE reads, P1 flow cell. Sequence reads were aligned to the nearest influenza A virus reference and sample consensus sequences were generated using IRMA [10]. Influenza clades were assigned based on submitting hemagglutinin sequences to Nextclade [11]. Sequences with ≥90% coverage of all 8 genomic segments were uploaded to GISAID and NCBI GenBank.

### US datasets

On October 1, 2025, we downloaded all influenza H3 and H1 hemagglutinin sequences from the U.S. for the time period starting on Oct 1, 2021, to Sept 30, 2025 (flu seasons 2021/22 to 24/25) from GISAID [12,13] and GenBank [14]. We deduplicated downloaded data based on isolate name, keeping a single sequence for each isolate (accession numbers and GISAID IDs are in Appendix, Table S1). The findings of this study are based on metadata associated with 45,194 sequences available on GISAID up to September 30, 2025, via gisaid.org/EPI_SET_251211sc, as well as 507 sequences from Genbank.

### Identifying mutating sites and circulating haplotypes

We used an augur workflow based on https://github.com/nextstrain/seasonal-flu to align and translate sequences [15]. Then, for each subtype and for each season, we used in-house scripts to identify sites with at least 2 amino acids with frequencies above pre-designated thresholds. We considered these sites our mutation sites, testing a range of thresholds: 0.005, 0.01, 0.05, 0.1, 0.15, 0.2. For example, a site identified at a threshold of 0.1 would have at least 2 amino acids, each at a frequency of 0.1 or above. Once we had identified sites with mutations, we generated haplotypes: the amino acid combinations across those sites, for each sequence. We defined haplotype frequencies for each season. We also calculated Shannon entropy across haplotypes for each season and subtype, using the equation below, where entropy H across all haplotypes X is calculated based on the frequency of each unique haplotype x:

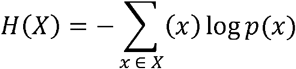

### Time to detection of haplotypes in Michigan

For each haplotype, we identified its first date of detection (which may have occurred earlier than the season in which the haplotype became dominant) and the date of its first detection in Michigan. We then fit two models to these data: a binomial logistic regression model predicting whether haplotypes were ever detected in Michigan, and a general linear model predicting the length of delays, for haplotypes that were detected in Michigan. Predictor variables tested were subtype, season, haplotype proportion (the proportion of circulating isolates represented by that haplotype), flu season week, haplotype length (i.e., number of variable sites), the number of days to haplotype extinction, seasonal entropy and the state where the haplotype was first detected. For each state we included additional state-specific metrics: state population size, distance to Michigan (state centroid to centroid), and state sequencing efforts (the number of sequences supplied per state and per season). Flu weeks were numbered from the first week in October (flu week 1) to the last week in September (flu week 52, or 53). Finally, we calculated epidemic proportion, the proportion of the final epidemic size that had already occurred when the haplotype was first detected using influenza-like illness (ILI) case counts and season totals obtained from ILInet [16]. As flu week and epidemic size were collinear, flu week was removed from the models. Because state was collinear with state metrics, we ran two version of the analyses, one where state was included as a variable, and one where state metrics were included. Statistical analyses were repeated across mutation thresholds.

### Downsampling analysis

To investigate how much UOM could decrease its sampling without negatively affecting haplotype detection delays, we carried out a downsampling analysis, downsampling Michigan sequences to 75%, 50%, 25% 10% and 5% of original dataset size and recalculating the numbers of undetected haplotypes and the delays to detection. Downsampling was repeated ten times at each downsample proportion, and results averaged across replicates. This analysis was performed at a single mutation threshold (0.1).

### Differences between states

Finally, to determine whether Michigan representativeness was higher than that of other states, irrespective of sample size, we downsampled the number of sequences from Michigan to the number of sequences from each state and compared how the state sample and the Michigan subsample compared in terms of number of undetected haplotypes, and delays to detection. This analysis was possible because the number of sequences from Michigan was greater than that from any other state in every season. Again, we downsampled ten times for each threshold, and analysis was performed at a single threshold for identifying mutations (0.1). We examined the relationship between sequencing effort and coverage through regression models.

To further investigate the extent of national diversity covered by state sequencing efforts, we conducted a rarefaction analysis. We constructed national haplotype frequency distribution, estimated the minimum number of sequences required to cover 95% of national diversity, and quantified sampling completeness for each state based on its sample sizes [17] using the iNext package in R [18].

All the code for conducting analyses was written in R and is available at https://github.com/manonr/Michigan_representativeness.git

## Results

### Michigan sequences account for 1/4 of sequences sampled in the U.S. from Oct 2021 onwards

Influenza seasons run from October 1^st^ to September 30^th^ of the following year. After deduplicating and data cleaning, our datasets comprised 19,282 H1 sequences: 82 from 2021/22, 2991 from 2022/23, 7437 from 2023/24, 8772 from 2024/25; and 26,419 H3 sequences: 6213 from 2021/22, 6110 from 2022/23, 5054 from 2023/24, 9042 from 2024/25 (Figure 1). Influenza sequencing across the U.S. has increased season on season since 2021, but Michigan sequenced more isolates in each of these four seasons (25.1% of the total number of sequences). Of the Michigan sequences in public databases, 95.7% were generated by us through various studies described in the Methods. Our analysis focuses on the representativeness of UOM sequences, which we refer to as the Michigan sequences. The 2021/22 and 2022/23 seasons were H3 predominant; 2023/24 and 2024/25 were mixed H3 and H1 flu seasons (Figure 1). The number of H1 sequences in the 2021 season was so small (n=82) that this subtype/ season was excluded from downstream analyses.

**Figure 1.**
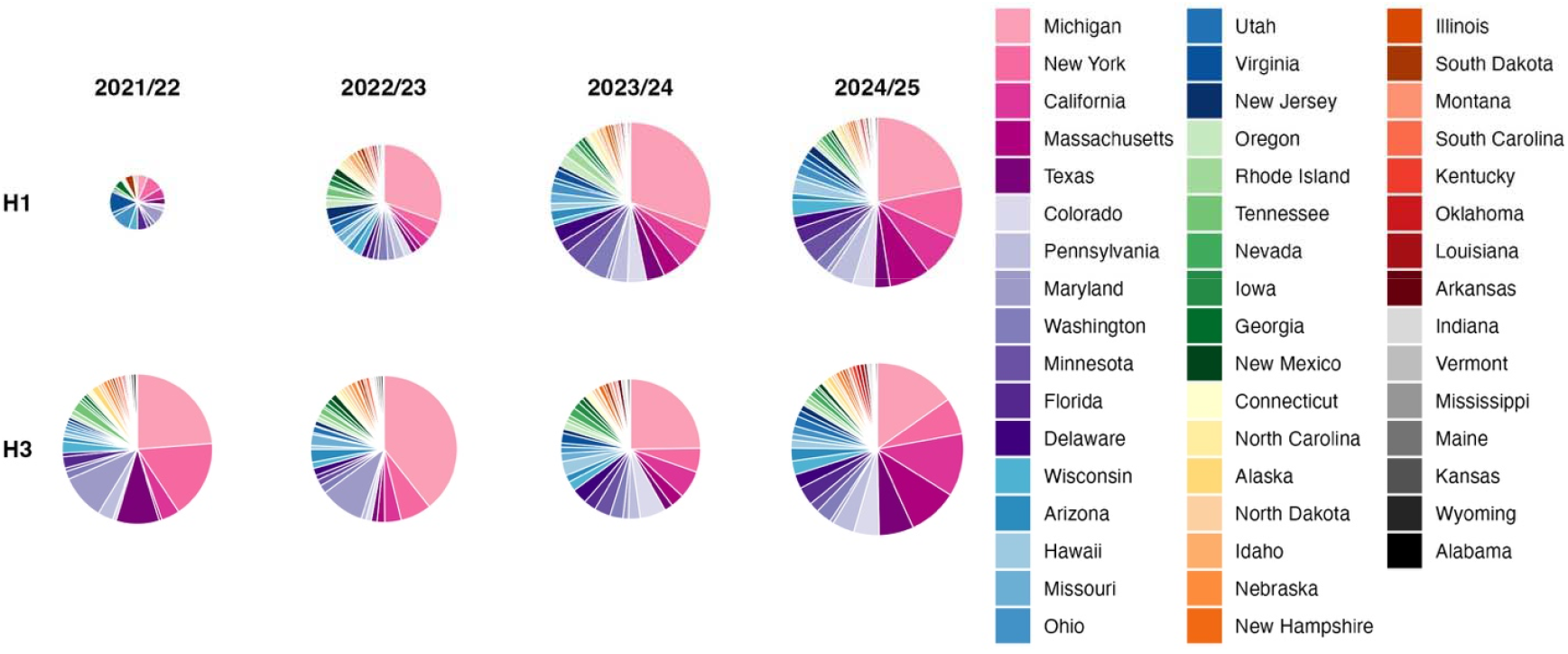
Number of publicly available sequences per season, state and subtype from 2021/22 to 2024/25. Influenza seasons run from October 1 to September 30th the following year. The size of circles is proportional to the overall number of sequences. States in the legend are ordered by overall number of isolates. Michigan sequenced more isolates than any other state in every season shown. Sequencing has increased overall during the study period.

### Seasonal diversity is determined by a small number of sites each season

The hemagglutinin segments analyzed here include 566 amino acid sites, for both H1 and H3. Across the entire dataset, most sites varied in at least one sequence: 446 for H3 and 468 for H1. However, when we focused on sites where mutation frequency was above a prespecified mutation frequency threshold (e.g., at least two different amino acids were present at that site above the threshold), the number of mutation sites dropped dramatically. We call the subset of sites that mutate above our pre-specified threshold within a season the seasonal “haplotype”.

At thresholds of 0.05 and above, the haplotype in a single season never exceeded 20 amino acid sites (Figure 2A), far fewer than the number of sites in defined structural epitopes. For H3, 129 sites are considered to be in epitopes [19], of which 121 varied within our dataset. For H1, 25 sites varied among the set of 31 structural epitopes [20]. At mutation thresholds below 0.05, the number of mutation sites increased dramatically, and we therefore limited downstream analyses to thresholds of ≥0.05 and their corresponding haplotypes.

**Figure 2.**
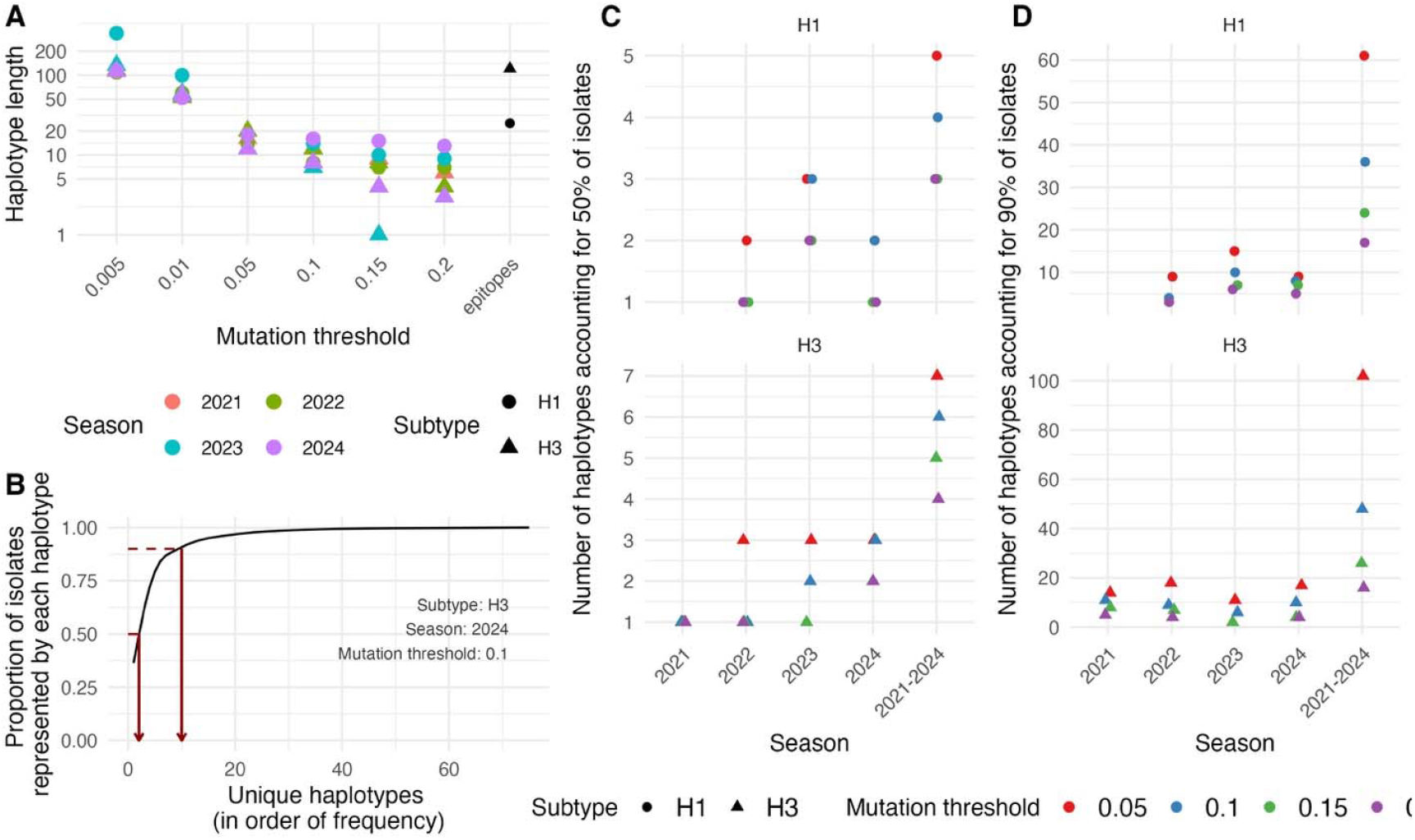
H3 and H1 haplotypes in the U.S. over four influenza seasons. (A) Haplotype length for different mutation thresholds. The number of total sites in structural epitopes for H1 (circle and H3 (triangle) are indicated. (B) Example haplotype cumulative distribution for a single subtype/ season/ threshold combination. (C) Number of haplotypes accounting for 50% of circulating isolates for H1 and H3, at different mutation thresholds. (D) Number of haplotypes accounting for 90% of circulating isolates for H1 and H3, at different mutation thresholds.

Based on the list of mutation sites in each season, we generated haplotypes for each sequence and season and calculated haplotype frequency for each season. The haplotype cumulative distribution had low variance, with a small number of haplotypes accounting for a large proportion of circulating isolates (Figure 2B). In fact, across all seasons, three or fewer haplotypes captured 50% of isolates (Figure 2C), and the number of haplotypes representing 90% of circulating isolates never exceeded 20 (Figure 2D). We observed more variation in H3 than in H1.

### The Michigan sample captures most strain diversity

For each haplotype, in order of frequency, we noted the first date of detection within the U.S. dataset, whether it was ever detected in Michigan, and if so, the date of detection. We present the analysis using a mutation threshold of 0.1. In every season and in both subtypes, >90% of U.S. isolates had a haplotype match in Michigan within a year (Figure 3A; Appendix, Figure S1 shows all delays and missing haplotypes), and the mean delay across seasons and subtypes was 107 days (SD=31). In most seasons, haplotypes representing the most frequent isolates are captured within the Michigan dataset within a few days of their first detection. Exceptions arise when that subtype accounts for a minority of circulating cases in that season, for example for H1 in 2022 (Figure 1, Figure 3).

**Figure 3.**
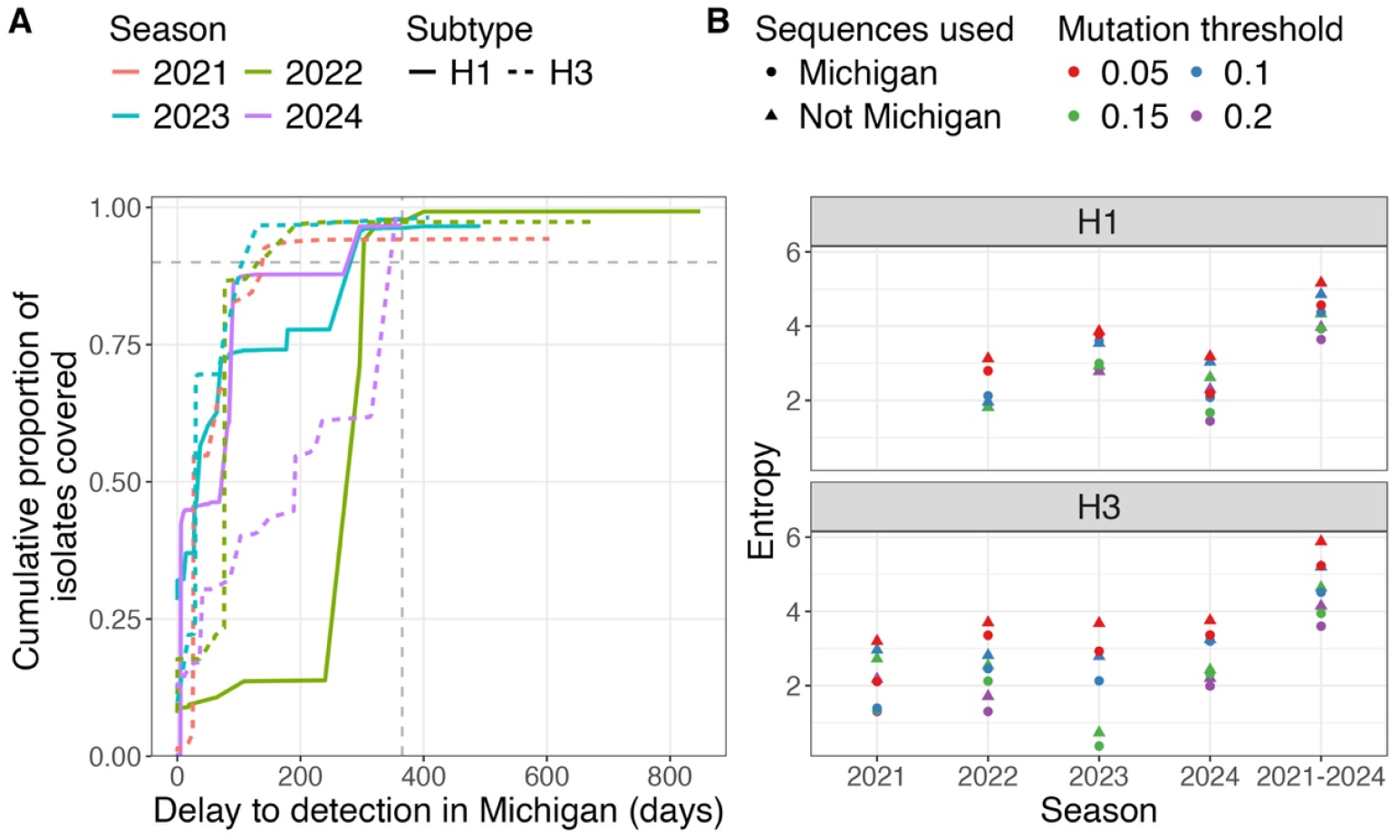
(A) Cumulative proportion of isolates captured by Michigan haplotypes, in days after the first appearance of the haplotype nationally, using a mutation threshold of 0.1. (B) Haplotype entropy in Michigan vs non-Michigan, for each season and each mutation threshold. The dashed lines in (A) indicate x=365 days and y=90%.

To evaluate the relative diversity within the Michigan dataset as compared to the rest of the U.S., we compared haplotype entropy between those two datasets. The entropy within Michigan and across the U.S. was similar, and entropy was relatively constant across seasons (Figure 3B), demonstrating that U.S. diversity is captured within Michigan.

### Causes of detection delays

Next, we examined the causes of delays or for haplotypes not being detected at all, to determine whether representativeness could be improved. We set up two regression models: a logistic regression model with a true/false outcome of whether a haplotype was ever detected in Michigan and a general linear regression model predicting the length of the delay. In both analyses we excluded haplotypes first detected in Michigan (n=58, out of 539 total haplotypes at a 0.1 mutation threshold). Again, we present results at a mutation threshold of 0.1, but results were entirely consistent across thresholds tested (0.05 to 0.2). Both higher haplotype proportion (OR>10^10^, p<0.005) and more days to haplotype extinction (OR=6.13, p<0.001) increased the probability of detection. A larger number of days to haplotype extinction was predictive of a longer delay to detection (β=51.60, p<0.001).

### Effect of downsampling

We investigated how much Michigan (or any other state conducting substantial sequencing) could reduce sequencing while maintaining coverage. We downsampled the Michigan dataset to 75%, 50%, 25%, 10% and 5% of its initial size, and recalculated the number of undetected haplotypes and the length of delays to detection. We present results at a mutation threshold of 0.1, with ten replicates for each downsample proportion.

Because a small number of haplotypes are representative of the large majority of circulating isolates, even with considerable downsampling, most circulating isolates had a haplotype match in Michigan (Figure 4A). However, delays to detection did increase with downsampling; when downsampling the Michigan dataset to only 5% of its original size, detection delays were doubled (Figure 4B).

**Figure 4.**
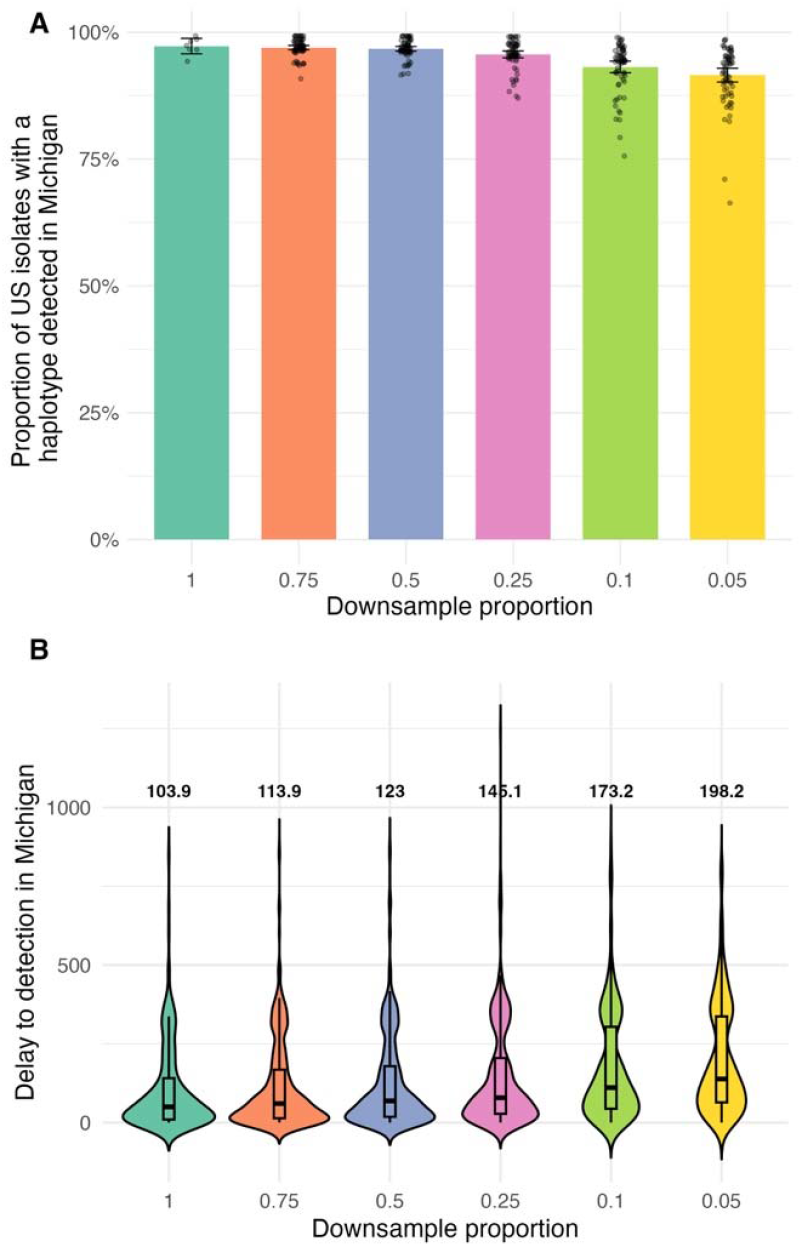
(A) Proportion of U.S. isolates with a Michigan haplotype match and (B) Delays to detection of haplotypes in Michigan, at various levels of downsampling (75%, 50%, 25%, 10%, 5%). In (A), the 95% confidence intervals (CI) represent the range across subtypes, seasons and replicates, and the result for each individual run is represented by a point. For the full sample, the intervals represent the 95% CI across subtypes and seasons. In (B), the numbers printed above each violin show the average delay to detection in Michigan.

### Comparison across states

Finally, we evaluated how haplotype coverage and detection delays in Michigan compared to those of other U.S. states. We identified haplotypes in the full U.S. dataset, and for each state constructed the national haplotype distribution excluding that state. We then measured the delay between the first appearance of each haplotype nationally and detection in that state, if it was detected at all.

To place each state’s performance in context, we also downsampled the Michigan dataset to match that state’s sample size. For each state, we generated a downsampled Michigan dataset of equivalent size and compared its haplotype coverage and detection delay metrics to the state’s own statistics. All sampling procedures were repeated across ten independent replicates. Then, for each state we generated an Area Under the Curve (AUC) metric, representing the proportion of isolates captured by a haplotype in that state within 365 days after the haplotype’s first detection (the curves would resemble those shown for Michigan in Figure 3). Our AUC metric thus represents both the proportion of circulating isolates captured by a haplotype in the focal state and speed to detection; lower AUC values reflect sparser coverage and slower detection.

Notably, patterns varied considerably from season to season (Figure 5). In 2022/23, where the H3 sample from Michigan was large as compared to other states, the Michigan samples do better overall than the individual states. By 2024/25, performance across states matched that of a Michigan downsample, largely because sample sizes had become more balanced. Where states perform significantly less well than the Michigan sample, this may be due to the non-randomness of their sampling strategy.

**Figure 5.**
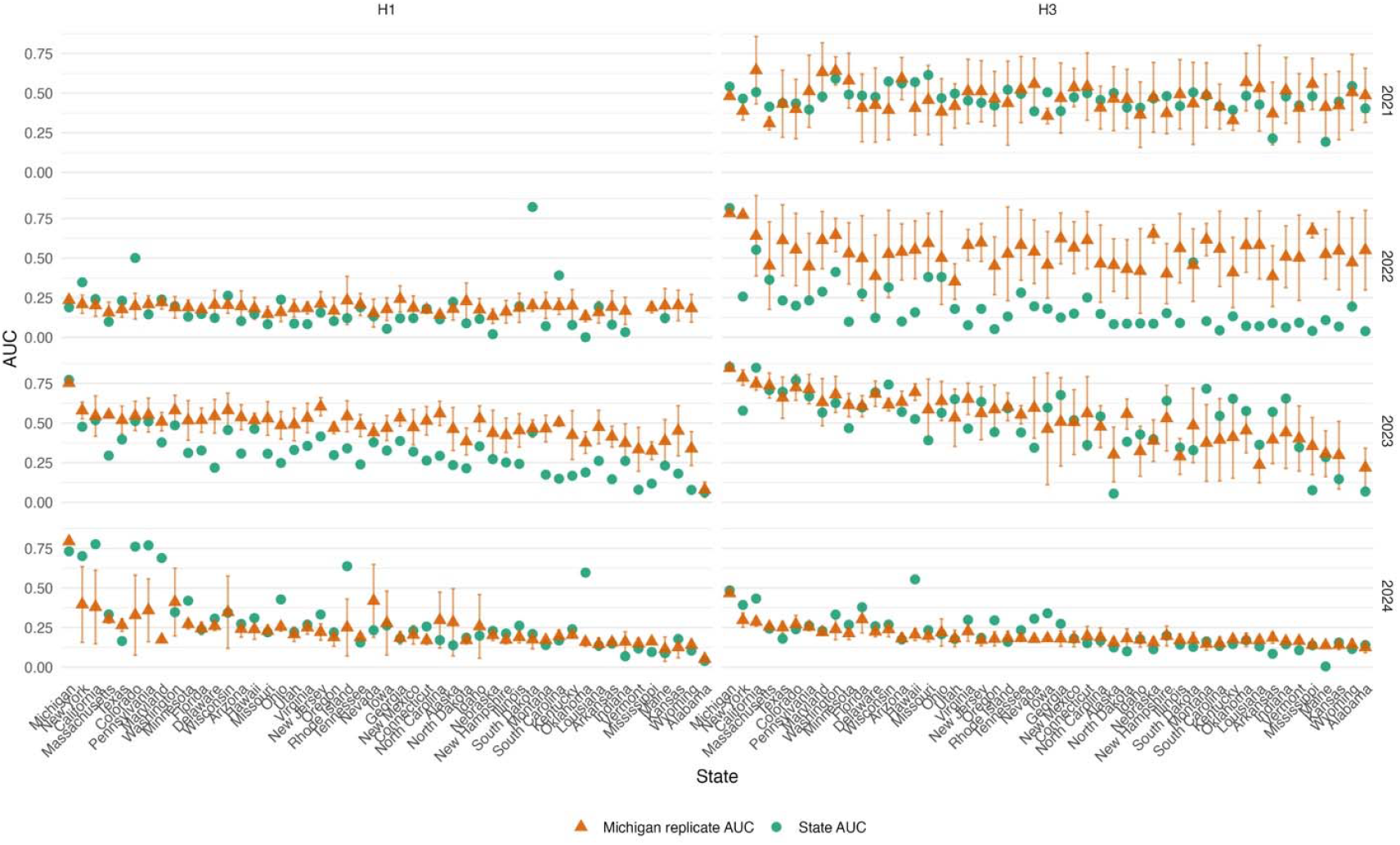
Coverage and time to detection are captured within the summary Area Under the Curve (AUC) metric. States are ordered by sequencing output across seasons and subtypes. For each state, the AUC is shown in green while the Michigan dataset sampled to the size of that state is shown in orange, with 95% indicating the range across ten replicates.

We further explored the relationship between sequencing effort and AUC using Gaussian regression models. Overall, AUC was positively associated with a log-transformed measure of state-level sequencing effort, consistent with diminishing returns as sequencing effort increased. Our final regression model, which included the log-transformed sequence count, season and subtype (Table 1), explained 52.4% of the variance in AUC. Season accounted for most of the variation in AUC, but there was no clear improvement in coverage between seasons 2021/22 and 2024/45. Including state and state interaction with sequence count increased the R^2^ to 69.1%; however, neither state nor its interaction reached individual significance in the model, indicating that the improvement was attributable to overfitting rather than systematic geographic variation. Including state population size as a covariate did not improve model fit This result is encouraging, as it suggests that despite non-random sampling, many states provide a broadly representative picture of circulating haplotypes so long as they sequence sufficiently.

**Table 1.** Predictors of AUC in regression model.

|  | Reference group | Estimate (95% CI) | p | Effect size |
| --- | --- | --- | --- | --- |
| Log number of sequences | NA | 0.07 ( 0.06, 0.09) | <0.001 | 0.29 |
| Season 2021/22 | Season 2024/25 | 0.22 ( 0.17, 0.27) | <0.001 | 0.37 |
| Season 2022/23 | Season 2024/25 | -0.03 (-0.07, 0.01) | NS |  |
| Season 2023/24 | Season 2024/25 | 0.18 ( 0.14, 0.22) | <0.001 |  |
| Subtype: H3 | Subtype: H1 | 0.05 ( 0.02, 0.08) | 0.004 | 0.03 |

Finally, the rarefaction analysis demonstrated that Michigan captured 99% of circulating national haplotype diversity in every season for both subtypes (other than H1 in 2021). Only two other states, California and New York, consistently exceeded 95% coverage (Appendix, Figure S2 and Table S2), despite the number of sequences required to cover 95% of haplotype diversity remaining small, reaching a maximum of n=227 for H1 in 2023.

## Discussion

Decisions based on influenza genomic surveillance are made at the national level, but data are acquired in a fragmented manner at state and local levels. We therefore evaluated the representativeness of these efforts. We found that a small number of haplotypes—defined here as sets of linked mutations among co-circulating strains—dominated each influenza season. Haplotype detection and delay to detection were primarily associated with national haplotype frequency and were not strongly affected by other factors examined, such as the state in which a haplotype was first detected. Accordingly, the number of sequences required to reach 95% coverage of national diversity was small across seasons and subtypes.

We examined whether sequencing from a single, well-sampled location could capture nationally circulating diversity. We found that all major haplotypes were detected in Michigan in every season, even with substantial downsampling. While Michigan performed better than other states in capturing this diversity, this advantage appeared to be mostly driven by higher sequencing volume rather than by sampling strategy. Together, these results suggest that intensive sequencing in a single U.S. location can yield data that are broadly representative of national haplotype diversity.

Comparisons across states demonstrated that higher sequencing effort was associated with more complete and rapid detection of circulating haplotypes. Heterogeneity across states was substantial and the best-performing state varied across seasons, consistent with dominant haplotypes emerging in different locations in different seasons [21]. After a certain level of sequencing coverage was achieved, there were no additional benefits from a random sequencing strategy such as that deployed in Michigan, but at lower rates of sampling, it improved diversity coverage. Only states with very low sequencing volumes performed consistently worse than a random sample.

Notably, our regression analyses searching for factors associated with detection delays or missed haplotypes yielded nothing beyond the strong association between detection and haplotype frequency, and detection and the number of days until haplotype extinction. No other variables examined - including subtype, season, epidemic stage, haplotype characteristics, the state of origin, and its sequencing effort - were with detection in Michigan. This finding again highlights the high probability of capturing high-frequency haplotypes, wherever one is sampling. In contrast, detection timing was more sensitive to sequencing effort. When fewer sequences were generated, the time required to detect emerging haplotypes increased sharply.

Han and colleagues, in their simulation study of SARS-CoV-2 genomic surveillance, concluded that the performance of surveillance systems for detecting emerging variants is constrained by access to diagnostic testing [22]. For detecting the dominant haplotype, no effect of diagnostic access was observed. Despite substantial variability in sequencing intensity across U.S. states, the dominant haplotypes were remarkably consistent nationwide, and Michigan continued to capture all major circulating haplotypes even under heavy downsampling. Recent work similarly concluded that few lineages in each U.S. influenza season accounted for more than half of circulating sequences (19).

Our study benefited from exceptionally dense sequencing, both in Michigan and nationally, owing to substantial federal investment in influenza genomic surveillance from 2021 to 2025 Even so, some states contributed very few sequences, and haplotypes could have circulated undetected in those locations. Our regression analyses did not indicate any trends associated with low-sequencing states, suggesting that undetected diversity would be minor, and unlikely to substantially alter our conclusions.

A potential criticism concerns our choice to define haplotypes rather than rely on existing clade systems or analyze individual mutations independently. Standard influenza clades are too coarse to capture seasonal dynamics and require dynamic naming that complicates retrospective comparison. At the opposite extreme, analyzing single mutations in isolation overlooks the epidemiologically and immunologically relevant genetic backgrounds on which influenza mutations arise. Our haplotype framework strikes a balance: it captures linked sets of seasonal co-circulating mutations, with the advantage of hindsight for classification, while preserving the genetic context that shapes viral evolution and antigenic change.

In conclusion, we found that Michigan influenza sequence data from 2021/22 to 2024/25 were highly representative of the viral diversity circulating across the U.S. Representativeness and detection timeliness improved during periods of more intensive sampling. Future work should continue to refine sampling strategies and assess how different approaches influence downstream analyses. It would also be valuable to investigate how inter-state differences in detection timing relate to the dynamics of regional epidemic across the U.S.

## Supporting information

Appendix, all supplementary figures + tables

## Data Availability

Sequences generated for this study were submitted to GISAID and Genbank. The findings of this study are based on metadata associated with 45,194 sequences
available on GISAID up to September 30, 2025, via gisaid.org/EPI_SET_251211sc and sequences on Genbank. All GISAID IDs and Accession numbers are detailed in Table S1.

https://doi.org/10.55876/gis8.251211sc

## Acknowledgements

We gratefully acknowledge all data contributors, i.e., the authors and their originating laboratories responsible for obtaining the specimens, and their submitting laboratories for generating the genetic sequence and metadata and sharing via the GISAID Initiative, on which this research is based. This work was supported by NIH U19 AI181767, NIH contract 75N93021C00015 (NIAID Centers of Excellence for Influenza Research and Response), and the Michigan Department of Health and Human Services (Michigan Sequencing Academic Partnerships for Public Health Innovation and Response).

## Biosketch first author

MRC is a Senior Research Scientist at the University of Chicago Department of Ecology & Evolution, Chicago.

## References

1. Huddleston J, Bedford T. Timely vaccine strain selection and genomic surveillance improve evolutionary forecast accuracy of seasonal influenza A/H3N2. eLife. 2025;14:RP104282. doi:10.7554/eLife.104282

2. Volz E, Mishra S, Chand M, et al. Assessing transmissibility of SARS-CoV-2 lineage B.1.1.7 in England. Nature. 2021;593(7858):266–269. doi:10.1038/s41586-021-03470-x

3. Figgins M, Lee J, Hadfield J, Bedford T. Nextstrain SARS-CoV-2 Forecasts. Nextstrain. Accessed November 26, 2025. https://nextstrain.org/sars-cov-2/forecasts

4. Huddleston J, Chang J, Lee J, Steinberg P, Neher R, Bedford T. Seasonal Influenza Circulation Patterns and Projections for September 2025 to September 2026. Zenodo; 2025. doi:10.5281/ZENODO.17281059

5. Huddleston J, Bedford T. Timely vaccine strain selection and genomic surveillance improves evolutionary forecast accuracy of seasonal influenza A/H3N2. eLife. 2025;14. doi:10.7554/eLife.104282.2

6. Volz EM, Koelle K, Bedford T. Viral Phylodynamics. PLoS Comput Biol. 2013;9(3):e1002947. doi:10.1371/journal.pcbi.1002947

7. Müller NF, Wüthrich D, Goldman N, et al. Characterising the epidemic spread of influenza A/H3N2 within a city through phylogenetics. PLOS Pathog. 2020;16(11):e1008984. doi:10.1371/journal.ppat.1008984

8. Centers for Disease Control, Association of Public Health Laboratories. Influenza Virologic Surveillance Right Size Roadmap. 2022.

9. Zhou B, Donnelly ME, Scholes DT, et al. Single-reaction genomic amplification accelerates sequencing and vaccine production for classical and Swine origin human influenza a viruses. J Virol. 2009;83(19):10309–10313. doi:10.1128/JVI.01109-09

10. Shepard SS, Meno S, Bahl J, Wilson MM, Barnes J, Neuhaus E. Viral deep sequencing needs an adaptive approach: IRMA, the iterative refinement meta-assembler. BMC Genomics. 2016;17(1):708. doi:10.1186/s12864-016-3030-6

11. Aksamentov I, Roemer C, Hodcroft EB, Neher RA. Nextclade: clade assignment, mutation calling and quality control for viral genomes. J Open Source Softw. 2021;6(67):3773. doi:10.21105/joss.03773

12. GISAID - gisaid.org. Accessed November 24, 2025. https://gisaid.org/

13. Elbe S, Buckland-Merrett G. Data, disease and diplomacy: GISAID’s innovative contribution to global health. Glob Chall. 2017;1(1):33–46. doi:10.1002/gch2.1018

14. GenBank. Accessed November 24, 2025. https://www.ncbi.nlm.nih.gov/genbank/about/

15. Hadfield J, Megill C, Bell SM, et al. Nextstrain: real-time tracking of pathogen evolution. Bioinformatics. 2018;34(23):4121–4123. doi:10.1093/bioinformatics/bty407

16. ILINet State Activity Indicator Map. Accessed November 24, 2025. https://gis.cdc.gov/grasp/fluview/main.html

17. Chao A, Gotelli NJ, Hsieh TC, et al. Rarefaction and extrapolation with Hill numbers: a framework for sampling and estimation in species diversity studies. Ecol Monogr. 2014;84(1):45–67. doi:10.1890/13-0133.1

18. Hsieh TC, Ma KH, Chao A. iNEXT: an R package for rarefaction and extrapolation of species diversity (Hill numbers). Methods Ecol Evol. 2016;7(12):1451–1456. doi:10.1111/2041-210X.12613

19. Wolf YI, Viboud C, Holmes EC, Koonin EV, Lipman DJ. Long intervals of stasis punctuated by bursts of positive selection in the seasonal evolution of influenza A virus. Biol Direct. 2006;1:34.

20. Xu R, Eckiert DC, Krause JC, Hai R, Crowe Jr JE, Wilson IA. Structural Basis of Preexisting Immunity to the 2009 H1N1 Pandemic Influenza Virus. Science. 2010;328(5976):357–360.

21. de Jong SPJ, Conlan AJK, Han AX, Russell CA. Competition between transmission lineages mediated by human mobility shapes seasonal influenza epidemics in the US. Nat Commun. 2025;16(1):4605. doi:10.1038/s41467-025-59757-4

22. Han A, Toporowski A, Sacks J, et al. SARS-CoV-2 diagnostic testing rates determine the sensitivity of genomic surveillance programs. Nat Genet. 2023;55:26–33.

