## Appendix, all supplementary figures + tables for "The Representativeness of Regional Influenza Virus Genomic Surveillance for National Trends in the United States"

Supplementary Tables and Figures

### **Table S1**

Genbank and GISAID IDs, GISAID acknowledgments (in separate csv)

### **Table S2**

#### Estimated coverage of national diversity for each state, season and subtype based on rarefaction analysis

The last column is the mean estimated coverage across subtypes and seasons, excluding H1 in 2021, and states are sorted by that mean.

|  | Estimated coverage of national diversity (estimated) | | | | | | | | |
| --- | --- | --- | --- | --- | --- | --- | --- | --- | --- |
| State | H1 | | | | H3 | | | | mean |
| Michigan | 0.39 | 0.99 | 0.99 | 0.99 | 0.99 | 0.99 | 0.99 | 0.99 | 0.99 |
| California | 0.39 | 0.96 | 0.97 | 0.96 | 0.96 | 0.95 | 0.99 | 0.98 | 0.97 |
| New York | 0.53 | 0.95 | 0.97 | 0.97 | 0.97 | 0.98 | 0.98 | 0.98 | 0.97 |
| Texas | 0.19 | 0.95 | 0.96 | 0.92 | 0.94 | 0.98 | 0.98 | 0.96 | 0.96 |
| Washington | 0 | 0.96 | 0.97 | 0.93 | 0.96 | 0.92 | 0.97 | 0.96 | 0.95 |
| Pennsylvania | 0.18 | 0.95 | 0.96 | 0.91 | 0.96 | 0.95 | 0.97 | 0.97 | 0.95 |
| Florida | 0.39 | 0.94 | 0.96 | 0.91 | 0.94 | 0.94 | 0.97 | 0.96 | 0.95 |
| Colorado | 0.1 | 0.95 | 0.97 | 0.91 | 0.95 | 0.88 | 0.98 | 0.97 | 0.94 |
| Minnesota | 0.1 | 0.96 | 0.97 | 0.92 | 0.93 | 0.88 | 0.95 | 0.97 | 0.94 |
| Wisconsin | 0.33 | 0.92 | 0.96 | 0.92 | 0.96 | 0.93 | 0.96 | 0.96 | 0.94 |
| Maryland | 0.49 | 0.9 | 0.95 | 0.98 | 0.95 | 0.98 | 0.91 | 0.93 | 0.94 |
| Massachusetts | NA | 0.95 | 0.97 | 0.93 | 0.94 | 0.86 | 0.98 | 0.97 | 0.94 |
| Delaware | NA | 0.95 | 0.97 | 0.93 | 0.94 | 0.88 | 0.96 | 0.96 | 0.94 |
| Arizona | NA | 0.94 | 0.95 | 0.95 | 0.95 | 0.9 | 0.96 | 0.94 | 0.94 |
| New Jersey | 0.1 | 0.91 | 0.94 | 0.91 | 0.96 | 0.87 | 0.94 | 0.95 | 0.93 |
| Utah | 0.18 | 0.91 | 0.94 | 0.92 | 0.96 | 0.86 | 0.95 | 0.94 | 0.93 |
| Hawaii | NA | 0.93 | 0.97 | 0.89 | 0.94 | 0.88 | 0.95 | 0.96 | 0.93 |
| Missouri | NA | 0.94 | 0.96 | 0.94 | 0.95 | 0.89 | 0.93 | 0.93 | 0.93 |
| Tennessee | 0 | 0.85 | 0.94 | 0.92 | 0.95 | 0.93 | 0.91 | 0.92 | 0.92 |
| Ohio | 0.54 | 0.94 | 0.95 | 0.84 | 0.91 | 0.87 | 0.95 | 0.95 | 0.92 |
| Oregon | NA | 0.94 | 0.95 | 0.9 | 0.95 | 0.87 | 0.9 | 0.91 | 0.92 |
| Iowa | NA | 0.91 | 0.95 | 0.9 | 0.94 | 0.87 | 0.91 | 0.93 | 0.92 |
| Rhode Island | NA | 0.94 | 0.93 | 0.9 | 0.93 | 0.9 | 0.93 | 0.92 | 0.92 |
| Virginia | 0.6 | 0.94 | 0.96 | 0.76 | 0.94 | 0.87 | 0.94 | 0.94 | 0.91 |
| Connecticut | 0.1 | 0.9 | 0.95 | 0.89 | 0.93 | 0.88 | 0.86 | 0.9 | 0.9 |
| Georgia | 0.19 | 0.9 | 0.95 | 0.91 | 0.94 | 0.88 | 0.85 | 0.9 | 0.9 |
| New Mexico | NA | 0.81 | 0.93 | 0.92 | 0.95 | 0.83 | 0.92 | 0.91 | 0.9 |
| Nevada | NA | 0.91 | 0.96 | 0.85 | 0.88 | 0.86 | 0.94 | 0.93 | 0.9 |
| Idaho | NA | 0.9 | 0.93 | 0.9 | 0.94 | 0.86 | 0.87 | 0.9 | 0.9 |
| North Dakota | NA | 0.88 | 0.92 | 0.91 | 0.92 | 0.88 | 0.9 | 0.9 | 0.9 |
| North Carolina | 0 | 0.91 | 0.86 | 0.89 | 0.91 | 0.83 | 0.92 | 0.92 | 0.89 |
| South Dakota | 0.33 | 0.86 | 0.94 | 0.89 | 0.93 | 0.86 | 0.87 | 0.87 | 0.89 |
| Nebraska | NA | 0.84 | 0.82 | 0.91 | 0.92 | 0.89 | 0.92 | 0.91 | 0.89 |
| Illinois | NA | 0.87 | 0.91 | 0.89 | 0.92 | 0.85 | 0.9 | 0.9 | 0.89 |
| New Hampshire | NA | 0.9 | 0.94 | 0.81 | 0.81 | 0.87 | 0.9 | 0.91 | 0.88 |
| Montana | 0.1 | 0.83 | 0.92 | 0.87 | 0.92 | 0.87 | 0.85 | 0.86 | 0.87 |
| South Carolina | NA | 0.84 | 0.79 | 0.89 | 0.88 | 0.87 | 0.9 | 0.87 | 0.86 |
| Oklahoma | NA | 0.78 | 0.86 | 0.79 | 0.91 | 0.81 | 0.92 | 0.87 | 0.85 |
| Alaska | NA | 0.72 | 0.58 | 0.88 | 0.92 | 0.91 | 0.92 | 0.92 | 0.84 |
| Kentucky | NA | 0.82 | 0.9 | 0.76 | 0.89 | 0.72 | 0.91 | 0.91 | 0.84 |
| Louisiana | NA | 0.84 | 0.69 | 0.79 | 0.89 | 0.83 | 0.91 | 0.84 | 0.83 |
| Indiana | NA | 0.77 | 0.93 | 0.77 | 0.72 | 0.81 | 0.88 | 0.85 | 0.82 |
| Mississippi | 0.1 | 0.69 | 0.74 | 0.85 | 0.89 | 0.81 | 0.82 | 0.84 | 0.81 |
| Maine | NA | 0.82 | 0.91 | 0.86 | 0.86 | 0.66 | 0.72 | 0.75 | 0.8 |
| Arkansas | NA | 0.79 | 0.93 | 0.47 | 0.72 | 0.77 | 0.89 | 0.89 | 0.78 |
| Vermont | NA | 0.69 | 0.84 | 0.84 | 0.57 | 0.76 | 0.87 | 0.9 | 0.78 |
| Kansas | NA | 0.69 | 0.72 | 0.86 | 0.83 | 0.85 | 0.72 | 0.75 | 0.77 |
| Wyoming | NA | 0.14 | NA | 0.74 | 0.8 | 0.86 | 0.81 | 0.84 | 0.7 |
| Alabama | NA | 0.51 | 0.41 | 0.47 | NA | 0.78 | 0.83 | 0.87 | 0.64 |

### Table S3

##### Number of sequences required to cover 95% of haplotype diversity, for each season and subtype.

The numbers are derived from the haplotype frequency distribution.

| Subtype | Season | Threshold | Total number of sequences | Number of sequences to cover 95% of haplotype diversity |
| --- | --- | --- | --- | --- |
| H1 | 2021 | 0.1 | 82 | 80 |
| H3 | 2021 | 0.1 | 6213 | 218 |
| H1 | 2022 | 0.1 | 2991 | 62 |
| H3 | 2022 | 0.1 | 6110 | 176 |
| H1 | 2023 | 0.1 | 7437 | 227 |
| H3 | 2023 | 0.1 | 5054 | 69 |
| H1 | 2024 | 0.1 | 8772 | 148 |
| H3 | 2024 | 0.1 | 9042 | 148 |

Figure S1
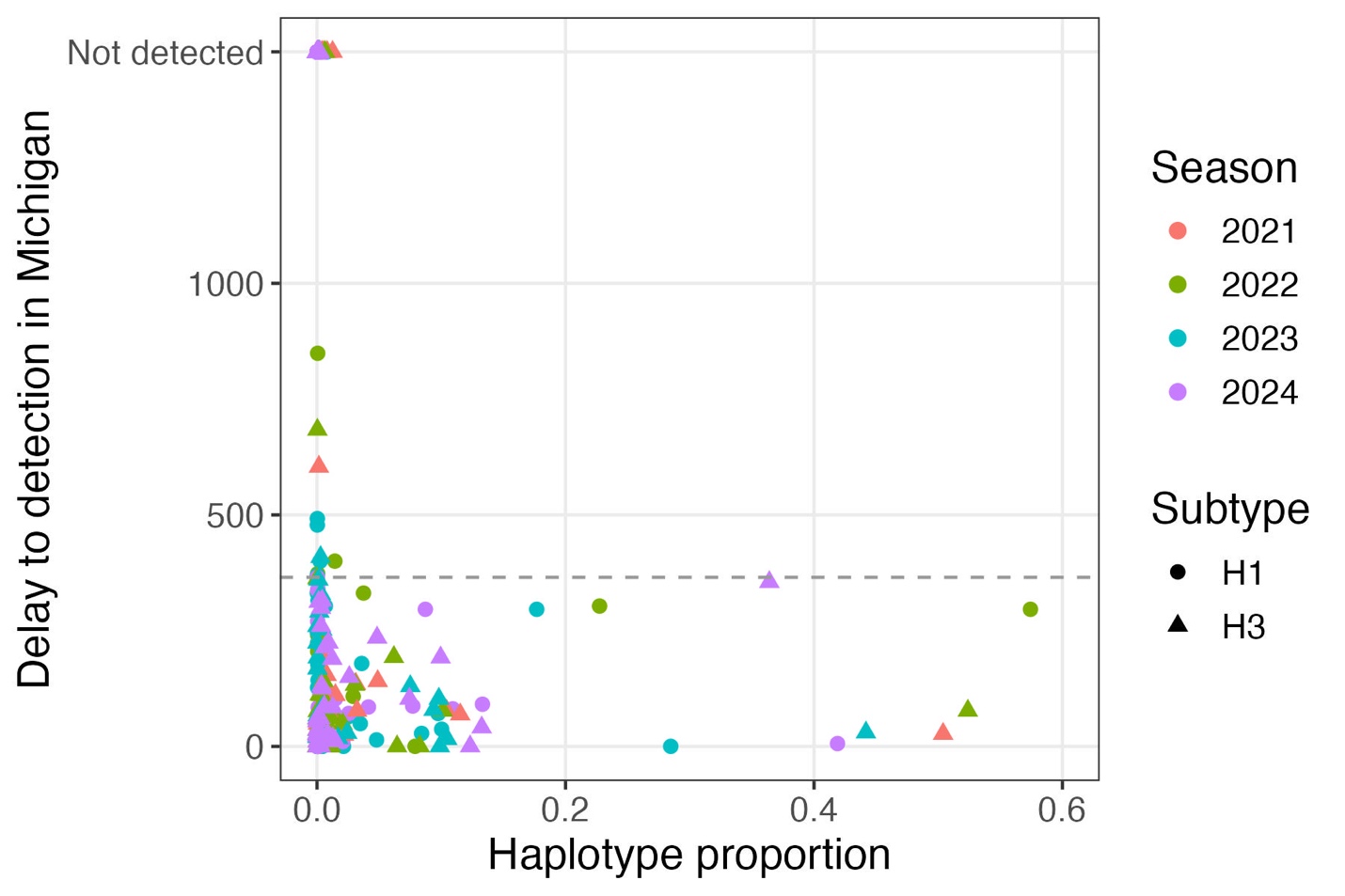

Supplementary Figure 1: Delay from first appearance nationally to detection in Michigan, according to haplotype proportion (the proportion of circulating isolates captured by that haplotype). The figure shows haplotypes defined at a mutation threshold of 1%. All haplotypes representing >5% of circulating were detected within less than 365 days. In every season and in both subtypes, >90% of US isolates had a haplotype match in Michigan within a year.

Figure S2

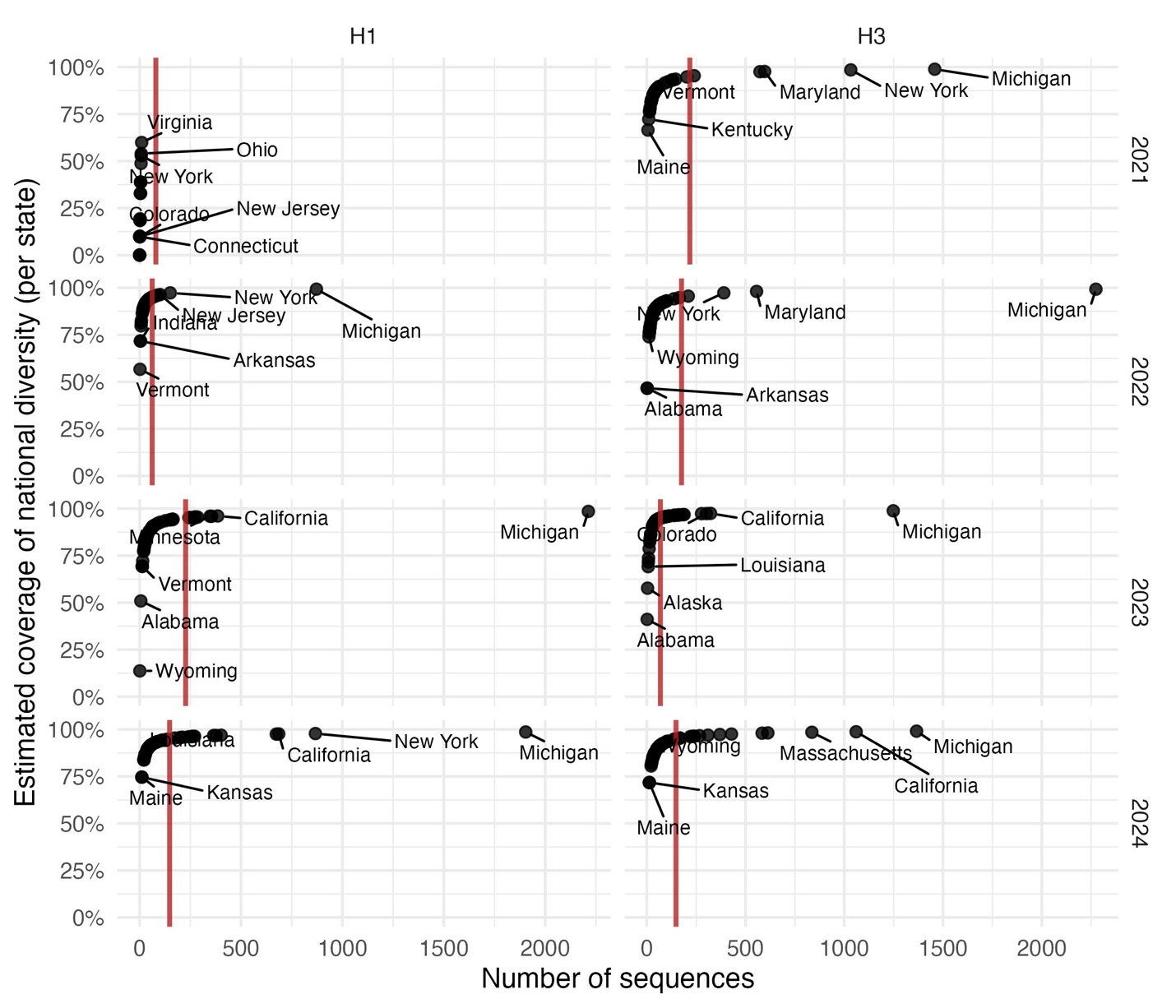

Supplementary Figure 2: Estimated coverage of national diversity for each state, season and subtype based on rarefaction analysis. The rarefaction analysis considers the national haplotype frequency distribution and state sample size. The red line represents the number of sequences required to cover 95% of national diversity. For each season and subtype, the three states with the highest and lowest coverage are annotated. Numerical details can be obtained in Table S2 and Table S3.
